# Organisms causing secondary pneumonias in COVID-19 patients at 5 UK ICUs as detected with the FilmArray test

**DOI:** 10.1101/2020.06.22.20131573

**Authors:** Zaneeta Dhesi, Virve I Enne, David Brealey, David M Livermore, Juliet High, Charlotte Russell, Antony Colles, Hala Kandil, Damien Mack, Daniel Martin, Valerie Page, Robert Parker, Kerry Roulston, Suveer Singh, Emmanuel Wey, Ann Marie Swart, Susan Stirling, Julie A Barber, Justin O’Grady, Vanya Gant

## Abstract

**Introduction:** Several viral respiratory infections - notably influenza - are associated with secondary bacterial infection and additional pathology. The extent to which this applies for COVID-19 is unknown. Accordingly, we aimed to define the bacteria causing secondary pneumonias in COVID-19 ICU patients using the FilmArray Pneumonia Panel, and to determine this test’s potential in COVID-19 management.

**Methods:** COVID-19 ICU patients with clinically-suspected secondary infection at 5 UK hospitals were tested with the FilmArray at point of care. We collected patient demographic data and compared FilmArray results with routine culture.

**Results:** We report results of 110 FilmArray tests on 94 patients (16 had 2 tests): 69 patients (73%) were male, the median age was 59 yrs; 92 were ventilated. Median hospital stay before testing was 14 days (range 1-38). Fifty-nine (54%) tests were positive, with 141 bacteria detected. Most were Enterobacterales (n=55, including *Klebsiella* spp. [n= 35]) or *Staphylococcus aureus* (n=13), as is typical of hospital and ventilator pneumonia. Community pathogens, including *Haemophilus influenzae* (n=8) and *Streptococcus pneumoniae* (n=1), were rarer. FilmArray detected one additional virus (Rhinovirus/Enterovirus) and no atypical bacteria. Fewer samples (28 % vs. 54%) were positive by routine culture, and fewer species were reported per sample; *Klebsiella* species remained the most prevalent pathogens.

**Conclusion:** FilmArray had a higher diagnostic yield than culture for ICU COVID-19 patients with suspected secondary pneumonias. The bacteria found mostly were Enterobacterales, *S. aureus* and *P. aeruginosa*, as in typical HAP/VAP, but with *Klebsiella* spp. more prominent. We found almost no viral co-infection. Turnaround from sample to results is around 1h 15 min compared with the usual 72h for culture, giving prescribers earlier data to inform antimicrobial decisions.

## INTRODUCTION

The emergence of SARS-CoV2 as a pandemic virus of global importance drives a need for clinical and pathological evidence upon which to base optimal therapeutic decisions. Whilst purely viral infections should not be treated with antibiotics, several respiratory viruses, notably influenza, are associated with secondary bacterial infection and additional pathology. These secondary infections reflect a combination of damage to the protective mucosa, facilitating bacterial colonisation and invasion, as well as virally-induced immunosuppression (1, 2). Viral and bacterial respiratory co-infections exacerbate disease severity, and can prompt ICU admission (3).

The extent to which COVID-19, as the disease caused by SARS-CoV2, is associated with secondary bacterial infection of the respiratory tract is unknown (4). Anecdotal evidence suggests that hospitalised COVID-19 patients are frequently prescribed empirical antimicrobials. Whether this is microbiologically necessary, even in severe cases, is unknown (5). In a brief review of existing literature Rawson *et al* conclude that there currently are insufficient data to inform empiric or reactive antibiotic decisions in a reasonable timeframe for critically-ill COVID-19 patients (5).

Irrespective of COVD-19, investigation of clinically-suspected bacterial pneumonia is complicated by the poor sensitivity of sputum culture and by the considerable interval (typically *circa* 72h) from sample to susceptibility test results. Recently-developed rapid tests have the potential to improve both the speed and sensitivity of investigation (6). INHALE (ISRCTN16483855) is a UK NIHR-funded research programme investigating the utility of rapid molecular diagnostics for the microbiological investigation of HAP/VAP in critical care (7). The programme incorporates an RCT, run across 12 UK hospitals, in which ICU patients with suspected hospital-acquired or ventilator-associated pneumonia (HAP/VAP) are randomised to have either (a) standard empirical therapy or (b) to have the BioFire FilmArray Pneumonia Panel test (bioMérieux) to support early treatment decisions (8). All patients have conventional microbiological investigation performed. The FilmArray is a PCR-based test with a turnaround time of 1h15min and can be performed inside the ICU. An antibiotic-prescribing algorithm is provided to support decision-making based on the results, thus going beyond a “Point of Care Test (POCT)” to provide ‘Point of Decision’ clinical support.

The COVID-19 pandemic resulted in recruitment to the INHALE trial being paused and, under the exigencies of the circumstances, we developed an observational sub-study to investigate the utility of the FilmArray Pneumonia Panel for the diagnosis and characterisation of secondary bacterial infection in COVID-19 ICU patients. Here, we report the results of this sub-study for 94 patients from 5 UK ICUs. The aims were to describe secondary bacterial, viral and “atypical” pathogens in COVID-19 ICU patients and to evaluate the potential of this panel for management of these patients.

## METHODS

Five adult ICUs participated: Aintree University Hospital (part of Liverpool University Hospitals NHS Foundation Trust), Chelsea and Westminster Hospital NHS Foundation Trust, Royal Free NHS Foundation Trust, University College London NHS Foundation Trust and Watford General Hospital (part of West Hertfordshire NHS Trust). The BioFire FilmArray Pneumonia Panel – seeking 18 bacterial, 9 viral, and 7 antibiotic resistance gene targets (9) – was run on the FilmArray Torch instrument, used as a POCT at or near the ICU. The test has a run time of 1h 15 min, with a loading time of approx. 2 min; utilisation followed the manufacturer’s instructions (10) with samples loaded by clinical ICU staff. The panel does not seek SARS-CoV2, and diagnosis of COVID-19 was based on separate testing by the hospitals.

Eligible cases had to be in-patients in a participating ICU and to have clinically-diagnosed, or PCR-proven COVID-19, with clinical features compatible with a suspected secondary bacterial pneumonia over and above those expected for “pure” COVID-19 viral pneumonia. They also needed to have sufficient surplus lower respiratory tract sample (200 μl sputum/ bronchoalveolar lavage [BAL] or endotracheal tube aspirate [ETA]) for testing on the FilmArray. In order to minimise COVID-19 infection risk, all FilmArray tests were performed in designated COVID-19 clinical areas by staff wearing full personal protective equipment suitable for invasive procedures, according to local guidelines. Test results were immediately delivered to the clinical ICU team along with the INHALE RCT prescribing algorithm (which allows some local variation), providing recommended treatment guidance (11). The foundation of this prescribing algorithm is to promote antimicrobial stewardship by indicating the narrowest spectrum antibiotic likely to cover the pathogen(s) detected and compatible with the patient’s penicillin allergy status. A second FilmArray test ≥ 5 days from the first Pneumonia Panel test was permitted if a new or continuing bacterial pneumonia was suspected. In parallel, a respiratory sample was sent to the hospital laboratory for routine microbiological investigation, performed according to the standard UK Laboratories Operating Procedures (12).

Baseline data including age, sex, comorbidities, date of COVID-19 diagnosis, admission to hospital, and ICU admission were collected. FilmArray test results and clinical microbiology results were recorded, along with a brief statement of the reason for FilmArray testing. Antibiotic prescribing data were also collected, but remain under analysis and will be reported separately. A bespoke REDCap database was used for data collection and storage (13); this provides a number of features to maintain data quality, including an audit trail, ability to query spurious data, search facilities, and validation of predefined parameters/missing data.

Chi Square tests were used to compare proportions.

Both the main trial and the sub-study have ethical approval from the London, Brighton and Sussex Research Ethics Committee (19/LO/0400) and the Health Research Authority.

## RESULTS

### Patient demographics

Up to cut-off date of this analysis (6 May, 2020), 98 patients had been recruited at the 5 ICUs (range 7-44 patients per site), with FilmArray results available for 94, all recruited after 3 April 2020. Sixteen patients had the test performed twice, giving a total of 110 reports. Demographic and background health data are summarised in Table 1.

**Table 1:**
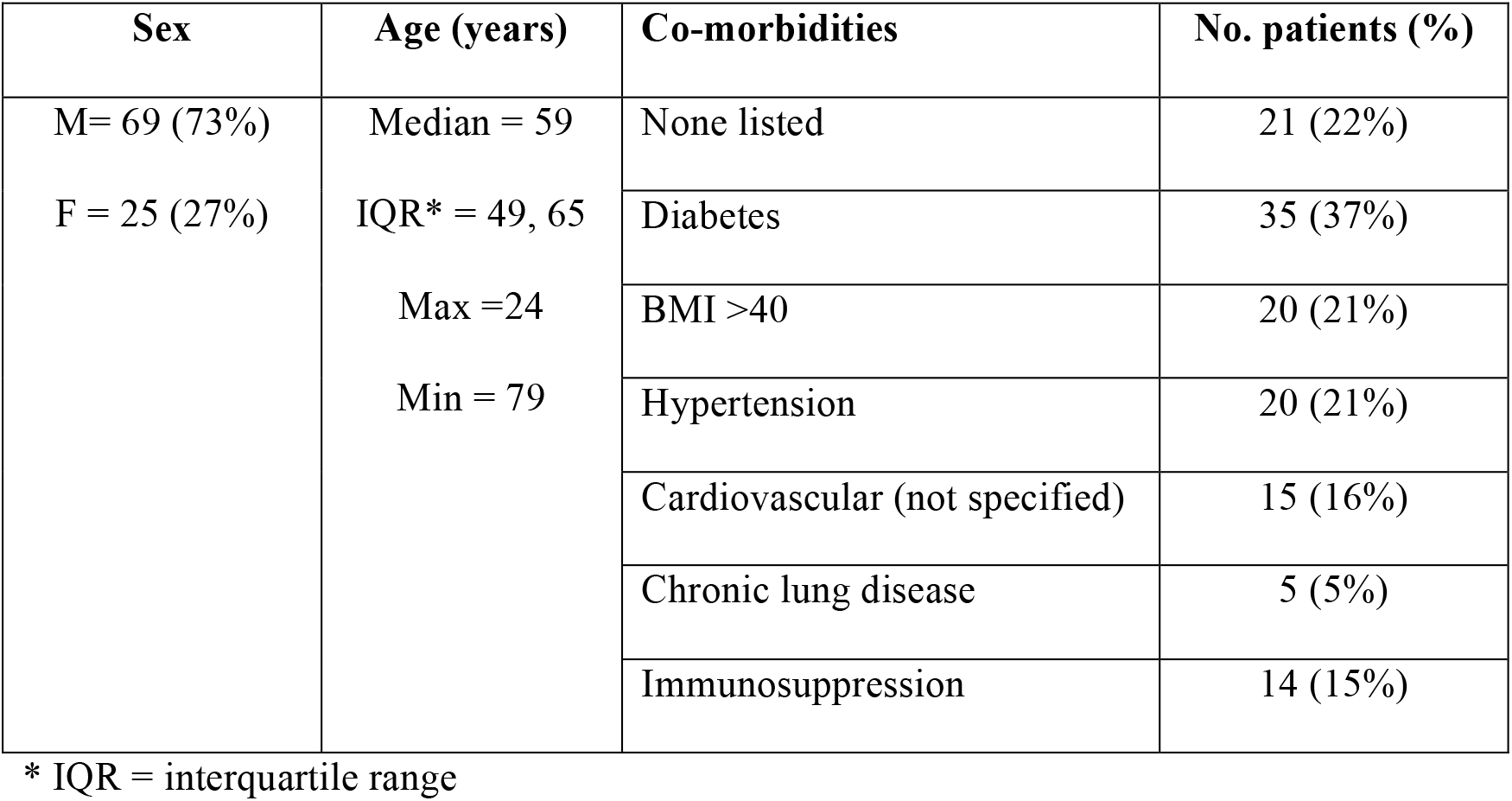
Demographics and co-morbidities of patients with FilmArray results (n = 94)

| Sex | Age (years) | Co-morbidities | No. patients (%) |
| --- | --- | --- | --- |
| M= 69 (73%)<br>F = 25 (27%) | Median = 59 | None listed | 21 (22%) |
|  | IQR* = 49, 65 | Diabetes | 35 (37%) |
|  | Max =24 | BMI >40 | 20 (21%) |
|  | Min = 79 | Hypertension | 20 (21%) |
|  |  | Cardiovascular (not specified) | 15 (16%) |
|  |  | Chronic lung disease | 5 (5%) |
|  |  | Immunosuppression | 14 (15%) |

\* IQR = interquartile range

Patients were hospitalised for a median of 14 days (range = 1-38 days; IQR 8,21) prior to FilmArray testing and were on an ICU for a median of 11 days (range = 0 - 37 days; IQR 7,17). All except two were ventilated; of the exceptions, one was breathing unassisted and the other was on non-invasive ventilation. Eighty-five patients had PCR-proven COVID-19 before FilmArray testing, 5 were PCR-negative but thought highly likely to have COVID-19 based on clinical presentation and/or imaging and 4 had COVID-19 tests pending at the time of FilmArray testing. Sample types used for FilmArray testing were 85 ETAs, 13 sputa, 5 non-directed BALs, 1 BAL, 1 ‘other’; 5 samples had no type specified in the database.

### Indication for FilmArray

Clinicians were asked to record the clinical indication for performing the FilmArray test. Among 69 responses provided, across the 5 participating sites, the most widely cited reason was an increase in inflammatory markers/fever (42%), followed by ‘considering a change of antibiotics’ (19%), suspected bacterial pneumonia (15%), increased respiratory secretions/ hypoxia (14%), or to stop antibiotics (10%).

### FilmArray results

Fifty-nine (54%) of the 110 tests proved positive for bacteria, whereas 51 (46%) had no organism(s) found. Among 6 FilmArray tests for patients who had been in hospital for <5 days, 3 were positive for bacteria: 2 contained *Staphylococcus aureus* only, while the third contained a mixture of Gram-negative species. Similarly, of 48 patients who had been in hospital for 15 days or more, 27 had a positive result. Among the 59 (54%) positive tests, 34 (58%) detected one organism only, 16 (27%) detected 2 organisms, 7 (12%) detected 3 organisms and 2 (3%) detected 4 organisms; a total of 90 non-replicate organisms (89 bacteria and 1 virus), plus 7 resistance gene sequences were recorded (Figure 1).

**Figure 1:**
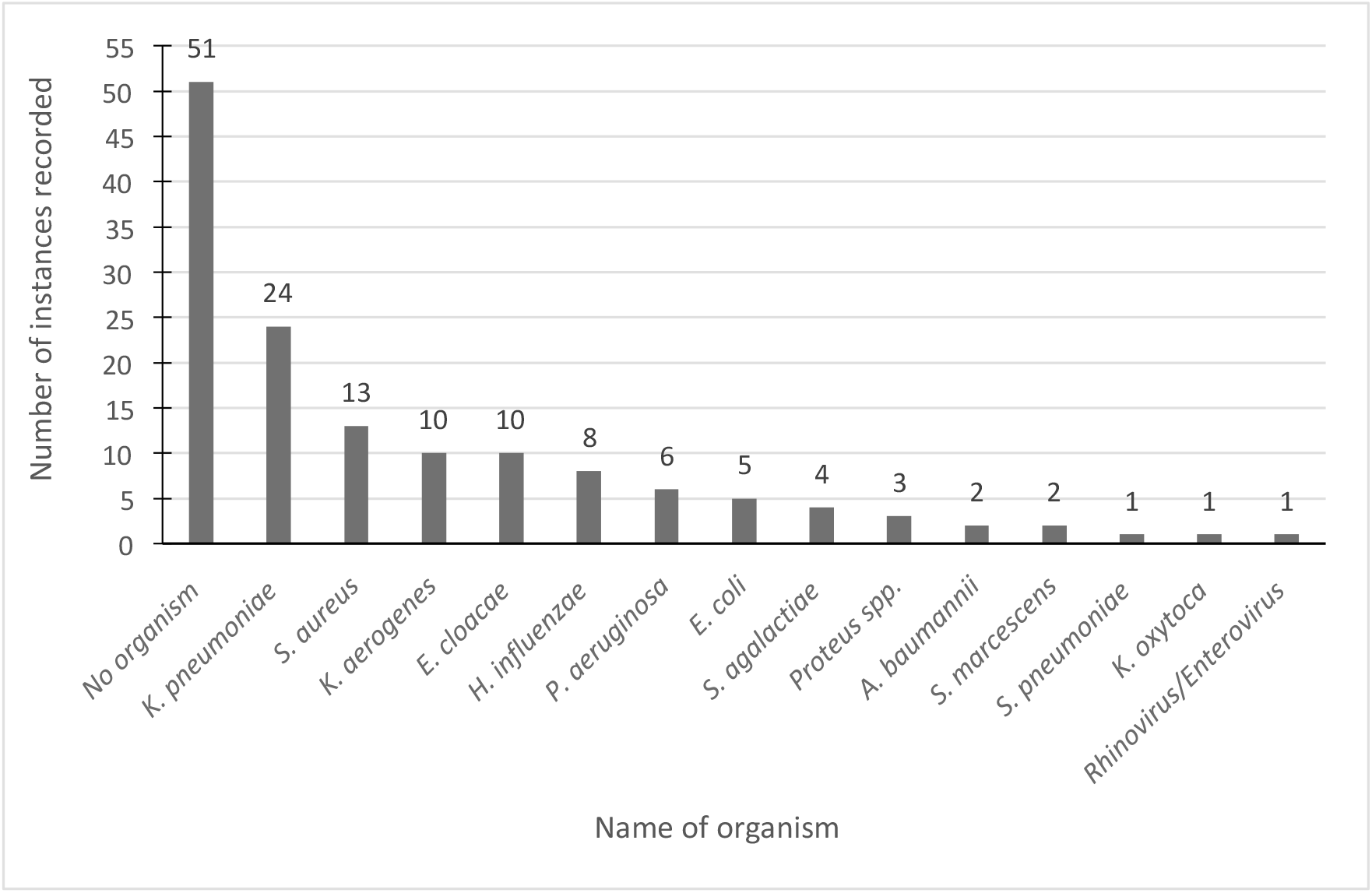
Organisms isolated by the FilmArray Pneumonia Panel (including negative results but excluding multiple instances of the same species from a single patient).

The most prevalent pathogen found was *Klebsiella pneumoniae*, followed by *S. aureus, Enterobacter cloacae* and *K. aerogenes*. Resistance genes were detected in 7 samples: *mec*A/C, which confers methicillin resistance in staphylococci, was found in 5 samples from 4 patients at 3 ICUs (one patient had two tests, with *mecA/C* found both times). All these source patients were positive for *S. aureus*, indicating an MRSA incidence of 4%. *bla*_CTX-M_ genes, encoding extended-spectrum β−lactamases, were detected in 2 samples, from different patients in the same ICU; both samples were positive for *K. pneumoniae*, a frequent host of these enzymes.

### Routine microbiology results

Out of the 110 specimens run on the FilmArray, 6 (5.5%) were not sent to the clinical microbiology laboratory and 5 (4.5%) were not reported; leaving 99 (90%) with a routine microbiology result (Figure 2). Two-thirds (66) of these 99 were reported as yielding ‘no growth/ significant growth’ or ‘normal respiratory flora’ with another 6 yielding only *Candida* spp., (2 more had *Candida* spp. plus another organism) which is not considered to be able to be an agent of pneumonia. This left 28 with bacteria reported. Multiple bacteria were reported from only 3 samples (2 organisms reported in 2 samples and 3 organisms reported in 1 sample), compared with 25/110 by FilmArray. Among the organisms reported, *K. pneumoniae* was again the most frequent, followed by *S. aureus*, as with the FilmArray.

**Figure 2:**
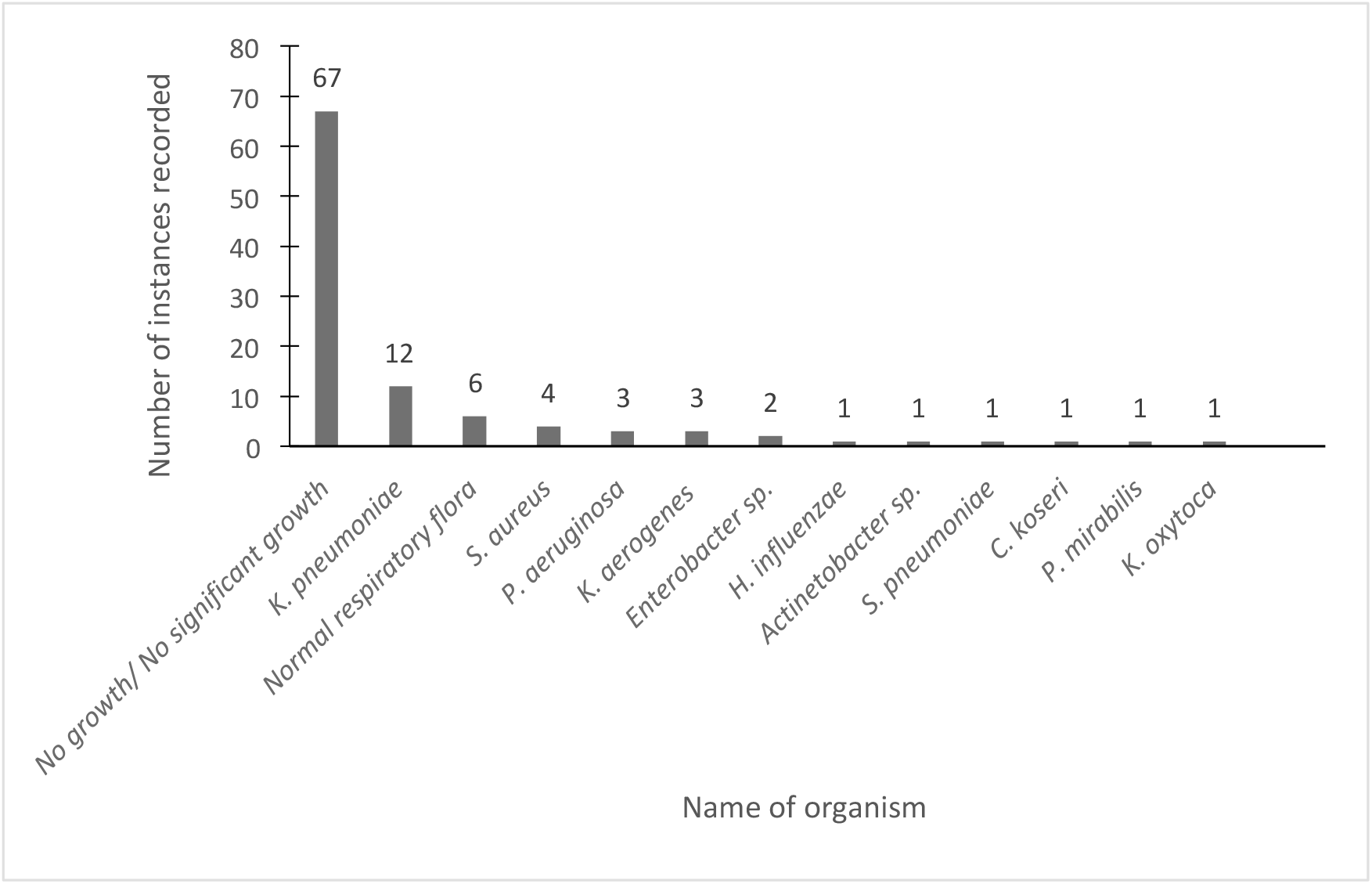
Bacteria reported by routine microbiology (including negative results but excluding multiple instances of the same species from a single patient).

### Comparison of FilmArray and routine microbiology

Three analyses were performed to compare the agreement of the FilmArray and routine results. First, we performed a concordance analysis, as shown in table 2, for tests where both results were available. Most FilmArray results (60.6%) were fully concordant with routine culture. Among those that were not fully concordant the common pattern was for FilmArray to indicate pathogens in samples where routine microbiology reported no organisms (25.3% of tests) or to flag additional organisms over and above those reported by routine microbiology (11.1%). Only 3% of cases were classified as major discordances with routine microbiology culturing an organism that was sought by the FilmArray but not found by it. One patient had an opportunist pathogen (*Citrobacter koseri*) not represented on the FilmArray panel.

**Table 2.** Concordance-based performance of FilmArray Pneumonia Panel compared with routine microbiology. N = 99 tests for which both results were available.

| Category* | Definition | Concordance,<br>No. samples (%)<br>(N=99) |
| --- | --- | --- |
| Full positive concordance | Organisms detected were an exact match | 14 (14.1) |
| Full negative concordance | No organisms detected by either method | 46 (46.5) |
| Partial concordance | FilmArray detected the same organism as routine microbiology plus additional organism(s) | 11 (11.1) |
| Minor discordance | Routine microbiology negative but FilmArray found $\geq 1$ organism* | 25 (25.3) |
| Major discordance | Routine microbiology found $\geq 1$ organism, at least one of which was on the FilmArray panel, but not detected | 3 (3.0) |

\*Includes one case where routine microbiology did not report *S. aureus* and *Klebsiella* spp., which were found by FilmArray, but did report *Citrobacter koseri*, which is not sought by FilmArray

Secondly, we reviewed agreement by species group in relation to the organism load reported by FilmArray (Table 3). Only 6 of 34 organisms reported by the FilmArray at a load of 10^4^ or 10^5^ CFU/ml were reported by routine microbiology, but this proportion rose to 19/44 for organisms found at a load of 10^6^ or 10^7^ CFU/ml (p=0.014, chi square test).

**Table 3:** Comparison of organisms detected by FilmArray at different concentrations and routine microbiology culture.

|  | Proportion of FilmArray detections where the same organism was reported by routine microbiology, among samples where at least one of the tests detected an organism |  |  |  |
| --- | --- | --- | --- | --- |
| Organism found by FilmArray | Reported by FilmArray<br>No estimate of microbial load) | Reported by FilmArray<br>(Load of 10<sup>4</sup> or 10<sup>5</sup> CFU/ml) | Reported by FilmArray<br>(Load of 10<sup>6</sup> or 10<sup>7</sup> CFU/ml) | Found by routine microbiology, not by FilmArray |
| *K. pneumoniae* |  | 3/11 | 7/12 | 1 |
| Other Enterobacterales |  | 1/13 | 4/13 | 2 |
| *H. influenzae* |  | 0/3 | 1/3 | 0 |
| *P. aeruginosa* & *A. baumannii* |  | 0/1 | 3/8 | 1 |
| *S. aureus* (all) |  | 2/6 | 3/7 | 0 |
| *S. aureus* and *mecA/C*<sup>a</sup> |  | 1/2 | 2/3 | 0 |
| *S. pneumoniae* |  | 0 | 1/1 | 0 |
| *Candida* spp.<sup>b</sup> |  |  |  | 8 |
| Rhinovirus/enterovirus<sup>c</sup> | 0/1 |  |  |  |
| Mycoplasma |  |  |  | 6<sup>d</sup> |

<sup>a</sup> Also included in 'All *S. aureus*' row
<sup>b</sup> Not sought by FilmArray
<sup>c</sup> Not sought by routine microbiology, routine virology data were not collected
<sup>d</sup> Found by serology, not confirmed by FilmArray PCR (data only available from one site)

Thirdly, we considered agreements between phenotypic resistance and detection of corresponding resistance genes by FilmArray. MRSA was reported by routine microbiology from 3 of the 5 samples where FilmArray flagged *mecA/C* and *S. aureus*; the remaining 2 samples were reported by microbiology as yielding ‘no growth’. Culture did not detect ESBL-producing organisms in either of the two samples where FilmArray found *bla*_CTX-M._ genes and *K. pneumoniae* but, curiously, did find an ESBL-producing *K. pneumoniae* in another sample (where FilmArray was negative) from the same ICU on the same day, raising a possible confusion of samples, though this could not be confirmed. Routine microbiology identified one *K. oxytoca* isolate with a phenotype suggesting hyper-production of K1 chromosomal β-lactamase; FilmArray detected the *K. oxytoca*, but does not seek the mutations that cause hyper-production of this enzyme.

There were just 3 cases of routine microbiology reporting organisms not detected by FilmArray: in one of these, FilmArray identified *S. marcescens* whereas microbiology isolated *K. aerogenes;* in another routine microbiology identified *Citrobacter koseri* (not on the Pneumonia Panel) whereas the FilmArray identified *K. pneumoniae*; and in the final case FilmArray identified a mixture of *H. influenzae, K. pneumoniae*, whereas routine microbiology identified both these organisms and *P. aeruginosa*.

One site found 6 patients were positive for either IgM or both IgM and IgG antibodies to *M. pneumoniae*: none of these were confirmed by the PCR test on the FilmArray; serological testing results have not yet been sought from other sites.

## DISCUSSION

To date, there are few data available regarding the occurrence and aetiology of secondary bacterial respiratory infections in COVID-19 patients. To redress this limitation, the present study examined these infections in severely-ill COVID-19 patients, all in ICU and almost all intubated. Patients were included based on suspicion of bacterial infection, consequently the results do not estimate the proportion of COVID-19 patients who develop secondary infection; rather, they illustrate the types of bacteria that are important in severe cases and the utility of the FilmArray PCR for detecting these. The age, sex and co-morbidity profiles of these patients are in keeping with those reported by others in severe COVID-19 disease (14).

Although the inclusion criteria permitted testing of patients at any point during their hospital and ICU admission, the majority (95%) of tests were performed at least 5 days after hospital admission. In context it should be noted that hospitalised COVID-19 patients in the UK are typically admitted to a general ward then, after several days, if necessary, are transferred to ICU, where our testing was conducted (14). Although sites had the option of using the test earlier during the hospitalisation, they reported anecdotally that most recently-hospitalised COVID-19 patients were unproductive for sputum and thus not eligible (data not shown). This observation may, of itself, suggest that early-onset bacterial secondary infections are uncommon in COVID-19 illness, as they would be expected to provoke sputum production. Nonetheless, it would be pertinent to examine the microbiology earlier in the patient’s hospital journey, e.g. on the day of admission, to see whether there were more negative results or if different organisms are isolated. However, the lack of sputum production may constrain such studies, especially where deliberate sampling, such as collecting BAL or induced sputum, is viewed as unnecessarily hazardous to staff and invasive for patients.

Among 110 FilmArray tests, representing 94 patients, 56% recorded bacteria or, in one case, a second virus. The microbiology found resembled that typical of HAP/VAP, in being dominated by Enterobacterales, *P. aeruginosa* and *S, aureus* (15). Organisms typically associated with community-acquired pneumonia were much less prominent: nevertheless *H. influenzae* was detected in 8 patients and *S. pneumoniae* in one, with all these detections relating to patients who had been hospitalised for at least 5 days, and for 22 days in the case of the *S. pneumoniae* patient. Similar observations in other studies of (COVID-19-unrelated) HAP suggest that these ‘community’ organisms can, on occasion, be hospital-acquired (15).

Despite the dominance of pathogens typically associated with HAP/VAP the species distribution differed from that seen in INHALE’s earlier evaluation study of the FilmArray test in HAP/VAP patients, conducted prior to the COVID-19 pandemic (16, 17). In particular, *Klebsiella* spp. (both *K. pneumoniae* and *K. aerogenes*) were significantly more prevalent (35/89 isolates *versus* 85/775, p <0.001; chi square test) in the COVID-19 patients, whereas *P. aeruginosa* and *E. coli* were under-represented, with the overall species distributions also significantly different (p <0.001; chi square test). An altered species distribution in HAP/VAP may reflect the particular thrombotic lung pathology associated with COVID-19 (18). The distribution of bacteria differed even more markedly from that typically seen following influenza, which is dominated by community-acquired pathogens such as *S. pneumoniae* and *H. influenzae*, with *S. aureus* also prominent (1). In China, Zhu *et al*. found *S. pneumoniae* to be the most prevalent bacterial pathogen in COVD-19 patients, followed by *K. pneumoniae* and *H. influenzae(19)*. In contrast to our study they mostly sampled early in the course of COVID-19 disease and, using throat swabs, examined patients who varied greatly in disease severity, meaning that comparability is tenuous. Also of note, only one of our 94 patients had an additional respiratory virus whereas 15.2% (94/620) of adult patients were positive for viruses in earlier INHALE work (16). This contrasts with data from China and California, where 22.6 - 31.5% of COVID-19 patients had co-infection with other viruses (19, 20). The key difference may be that we specifically examined ICU patients, many of whom had been hospitalised for prolonged periods, whereas these authors examined broader groups of COVID-19 patients with more recent community residency. Alternatively, the difference may be that these studies were done up to March 2020, and so overlapped the winter respiratory season, whereas we recruited later, in April and May.

In general, FilmArray identified a larger proportion of samples as positive for bacteria than routine culture (54 % vs. 28%); moreover, FilmArray more often indicated multiple bacteria in a sample. These findings accord with our previous observations, where we demonstrated that various PCR systems including FilmArray, Curetis Unyvero and also 16S rDNA analysis, all tended to find more organisms than are reported by routine culture from respiratory samples and that they tended also to find the same additional organisms as one another, implying that the vast majority of these additional detections represent organisms genuinely present in the sample (16).

A curious discrepancy was that routine serology, only obtained from one hospital, reported 6 cases positive for mycoplasma among the cohort (Table 3). *M. pneumoniae* was not detected by FilmArray PCR in the corresponding specimens, suggesting either that the serology represented a false positive result, perhaps owing to an anamnestic response, as seen with Dengue serology (21), or that mycoplasma species other than *M. pneumoniae* were present.

ICU clinicians have welcomed this new diagnostic platform to aid the rapid detection (or not) of bacteria in their patients’ lower respiratory tracts, and as a guide to treatment. The hazard is, however, that the greater diagnostic yield compared with culture may lead to treatment of patients who merely had a few colonising bacteria. The significance of organisms detected at low population densities (10^4^ to 10^5^ CFU/ml) remains open to debate; those found at higher densities were more often reported also by routine microbiology, with this differentiation stronger than in the main INHALE trial. More generally, we would underscore that the clinical context must be taken into account and that, as with many microbiological results, detection of an organism does not prove that it is causing infection. Balancing these factors will need careful liaison between ICUs, microbiology and other antimicrobial stewards; furthermore, clinical antibiotic prescribing decisions are subject to factors beyond a valid test result, as demonstrated in the VAPrapid study (22). That said, preliminary observational data from INHALE’s earlier work suggested that treatment of additional organisms detected by PCR may have the potential to improve patient outcomes (17). To examine the impact of FilmArray results in the context of COVID-19, we are collecting and analysing additional data at the 2 largest of the present 5 sites, assessing consequences for antimicrobial prescribing and patient outcomes. A behavioural sub-study is also underway to investigate how antibiotic decision making has been affected during the COVID-19 pandemic, and how this is influenced by the FilmArray.

In summary we have shown, first, that the bacteria causing secondary pneumonias in severely-ill COVID-19 patients mostly are Enterobacterales, *S. aureus* and *P. aeruginosa*, as is typical of HAP/VAP. The organism distribution is different from ‘typical’ HAP/VAP, with *K. pneumoniae* and *K. aerogenes* more prominent and *E. coli and P. aeruginosa* less prominent. Secondly, severe COVID-19 patients do not appear to progress to secondary bacterial infection in the same way as do severe influenza patients and do not have the same pathogens; rather, invasive ventilation seems likely to be the main driver for secondary infections in COVID-19. Thirdly, we have shown that FilmArray had a higher diagnostic yield than culture– as reported also in INHALE’s pre-COVID-19 work (16, 17). Turnaround from sample to results was around 1h 15 min compared with the usual 72h for culture, giving prescribers earlier data to inform antimicrobial decisions. Further work is required to establish the contribution of secondary infections to the overall clinical outcome in severely ill COVID-19 patients.

## Data Availability

Data available upon request.

## Acknowledgements

We thank all the ICU research nurses who have collected the data for this study, specifically: University College London Hospitals NHS Foundation Trust: Debbie Smyth and ICU research team. Royal Free Hospital: Helder Filipe and the ICU research team.

Chelsea and Westminster Hospital: Rhian Bull and the ICU research team.

Aintree University Hospital: Ian Turner Bone and the ICU research team.

Watford General Hospital: Xiao Bei Zhao and the ICU research team.

We thank bioMérieux for providing the machines and tests.

## Transparency Declarations

VE: has received speaking honoraria, consultancy fees and in-kind contributions from several diagnostic companies including Curetis GmbH, bioMérieux and Oxford Nanopore.

DB: has received speaking honoraria or consultancy fees from bioMérieux, Gilead and T2 Biosystems.

DML: Advisory Boards or ad-hoc consultancy Accelerate, Allecra, Antabio, Centauri, Entasis, GlaxoSmithKline, Meiji, Melinta, Menarini, Mutabilis, Nordic, ParaPharm, Pfizer, QPEX, Roche, Sandoz, Shionogi, T.A.Z., Tetraphase, Venatorx, Wockhardt, Zambon, Paid lectures –

Astellas, bioMérieux, Beckman Coulter, Cardiome, Cepheid, Merck/MSD, Menarini, Nordic, Pfizer and Shionogi. Relevant shareholdings or options – Dechra, GSK, Merck, Perkin Elmer, Pfizer, T.A.Z, amounting to <10% of portfolio value.

HK: Has received speaking and travel honoraria from bioMérieux.

DMa: Consultancy and lecture fees from Siemens Healthineers and Edwards Live Sciences.

VP: Has received honorarium and expenses from Orion pharma UK March 2017.

JOG: has received speaking honoraria, consultancy fees, in-kind contributions or research funding from Oxford Nanopore, Simcere, Becton-Dickinson and Heraeus Medical.

VG: has received speaking honoraria from BioMérieux and support for Conference attendances from Merck/MSD and Gilead.

ZD, JH, CR, AC, DM, RP, KR, SS, JB, AMS Nothing to declare.

## REFERENCES

1. Bakaletz LO. Viral-bacterial co-infections in the respiratory tract. Curr Opin Microbiol. 2017;35:30–5.

2. Rynda-Apple A, Robinson KM, Alcorn JF. Influenza and Bacterial Superinfection: Illuminating the Immunologic Mechanisms of Disease. Infect Immun. 2015;83:3764–70.

3. Cawcutt K, Kalil AC. Pneumonia with bacterial and viral coinfection. Curr Opin Crit Care. 2017;23:385–90.

4. Cox MJ, Loman N, Bogaert D, O’Grady J. Co-infections: potentially lethal and unexplored in COVID-19. The Lancet Microbe. 2020;1.

5. Rawson TM, Moore LSP, Gilchrist M, et al. Bacterial and fungal co-infection in individuals with coronavirus: A rapid review to support COVID-19 antimicrobial prescribing. Clin Infect Dis. 2020.

6. Poole S, Clark TW. Rapid syndromic molecular testing in pneumonia: The current landscape and future potential. J Infect. 2020;80:1–7.

7. INHALE TRIAL, UCL https://www.ucl.ac.uk/inhale-project/.

8. High J, et al. The Impact of using FilmArray Pneumonia Panel Molecluar Diagnostics for Hospital-Acquired and Ventilator-Associated Pneumonia on Antimicrobial Stewardship and Patient Outcomes in UK Critical Care: A Multicentre Randomised Controlled Trial. In preparation 2020.

9. bioMerieux. FilmArray Pneumonia Panel https://www.biofiredx.com/products/the-filmarray-panels/filmarray-pneumonia/.

10. bioMerieux. FilmArray Pneumonia Panel Instructions for Use EN. https://www.online-ifu.com/ITI0040/25003/EN

11. Dhesi Z, Enne VI, Gant V, Livermore D. Designing an Antibiotic Prescribing Algorithm to Complement Rapid Microbiological Investigation of Hospital-aquired and Ventilator-assoicated Pneumonia with the FilmArray Pneumonia Panel Plus: The INHALE Trial. In preparation 2020.

12. Public Health England. UK Standards for Microbiology Investigations. Investigation of bronchoalveolar lavage, sputum and associated specimens. https://assets.publishing.service.gov.uk/government/uploads/system/uploads/attachment_data/file/800451/B_57i3.5.pdf

13. Research Electronic data capture REDCap. https://www.project-redcap.org/ Accessed 01 June 2020. 2020.

14. ICNARC. Report on COVID-19 in critical care 12 June 2020.

15. Masterton RG, Galloway A, French G, et al. Guidelines for the management of hospital-acquired pneumonia in the UK: report of the working party on hospital-acquired pneumonia of the British Society for Antimicrobial Chemotherapy. J Antimicrob Chemother. 2008;62(1):5–34.

16. Enne VI, Aydin A, Richardson H, et al. Performance of two multiplex PCR platforms against routine microbiology for the detection of pathogens causing nosocomial pneumonia across 15 intensive care units in the UK. In preparation 2020.

17. Enne VI, Baldon R, Russell C, et al. INHALE WP2: Appropriateness of Antimicrobial Prescribing for Hospital-acquired and Ventilator-associated pneumonia in UK ICUs assessed aganist PCR-based molecluar diagnostic tests. 29th ECCMID 2019 Abstract 2019.

18. Gavriilaki E, Brodsky RA. Severe COVID-19 infection and thrombotic microangiopathy: success does not come easily. Br J Haematol. 2020;189:e227–e30.

19. Zhu X, Ge Y, Cui L, et al. Co-infection with respiratory pathogens among COVID-2019 cases. Virus Res. 2020;285:198005.

20. Stanford, Medicine, Data. Higher co-infection rates in COVID19. 2020. https://mediumcom/@nigam/higher-co-infection-rates-in-covid19-b24965088333.

21. Wang Q, Du Q, Guo X, et al. A method to prevent SARS-CoV-2 IgM false positives in gold immunochromatography and enzyme-linked immunosorbent assays. J Clin Microbiol. 2020.

22. Hellyer TP, McAuley DF, Simpson AJ, et al. Biomarker-guided antibiotic stewardship in suspected ventilator-associated pneumonia (VAPrapid2): a randomised controlled trial and process evaluation. Lancet Respir Med. 2020;8:182–91.

